# The Impact of Premature Birth and Low Birth Weight on Motor, Visual, and Cognitive Skills and Mental Health in Adolescence: A Systematic Review and Meta-analysis

**DOI:** 10.1101/2025.05.02.25326710

**Authors:** F. Schröpfer, E. Greif, S. Eickhoff, H. Schewe, J. Wiese, N. Derner, L. Heller, C. Andreou, K. Röse, J. Obleser, W. Göpel, S. Borgwardt, L. Franzen

## Abstract

Mental disorders represent a significant challenge for individuals and society. Many of them have a detectable onset during adolescence. Being born preterm or with low birth weight (PTB) has been emerging as a potential risk factor for developing mental health disorders in adolescence. Since PTB infants are also at an increased risk of developing cerebral visual impairment after birth and cognitive and sensory deficits throughout life, this systematic review seeks to understand the interplay between PTB, cognitive, visual, and motor abilities and their influence on mental health outcomes in adolescence.

We conducted a registered systematic review following the PRISMA guidelines (PROSPERO #513150). The search strategy focused on the databases PubMed, Scopus, PsycINFO and the Cochrane Library and included publications sampling participants born in 1980 or later.

We analysed 18 studies including a total of 13.655 adolescents aged 11-20 (PTB=8.813, control=4.852) published between 2004 and 2022. PTB adolescents exhibited consistent and persistent deficits in cognitive and motor domains compared to their full-term (FT) peers, including lower intelligent quotient (IQ), attention and executive function, and a higher prevalence of psychiatric disorders. Several studies demonstrate that PTB adolescents require psychiatric treatment more often and get diagnosed with more complex psychiatric disorders. Evidence for functional visual alterations is scarce.

These findings highlight the multitude of challenges that PTB children face across multiple domains including their mental health. Quantifiying these challenges individually may represent potential markers for tailoring early detection and intervention to this vulnerable population and optimising their long-term outcomes.

## Introduction

Mental well-being among adolescents in the WHO European Region declines with age and shows significant gender disparities: 15-year-old girls score lowest on the WHO-5 Well-being Index (49.9 vs. 62.0 for boys), report worse health (23% vs. 40%), and higher loneliness (28% vs. 13%) [1]. Mental health policies exist in most member states (86% for those under 19; 77% for those aged 15–24) [1]. With most mental disorders having a detectable onset around age 15, mental ill-health contributes to at least 45% of the disease burden in 10–24-year-olds and is often met with stigma and limited access to care—especially in low-resource settings, where services remain fragmented and data scarce [2].

As early as 2011, it was estimated that one-third of the EU population of all ages—around 165 million people—were affected by mental disorders annually [3]. The persistence and severity of these disorders have major consequences at both individual and societal levels. If a mental disorder begins in early adulthood, affected individuals may experience a reduction in life expectancy of 10–20 years [4]. It is during late adolescence and early adulthood that most mental disorders become clinically detectable [5–7]. Their first signs, including subtle cognitive, psychological, and physical changes, may already appear during prodromal phases up to five years prior, often during adolescence, as is the case with psychotic disorders [5].

Adolescence, defined by the WHO as ages 11 to 20 [6], marks a critical transition from childhood to adulthood. This period involves rapid biological, neural, and psychological changes [7, 8], leaving the window from conception to early adulthood for early detection and intervention.

Successful early detection of mental health problems heavily depends on the identification of the most informative risk factors. One risk factor may be being born preterm or with low birth weight (PTB). PTB is increasingly recognised as a key factor that may influence later neurodevelopmental outcomes and mental well-being [8]. Infants born extremely preterm (<26 weeks of gestation) or with extremely low birth weight (<1,000 g) face a 3-4 fold risk of developing a mental disorder later in life [9]. For example, schizophrenia is likely caused by an interplay of genetic and environmental factors that may include pre- and perinatal adversity, suboptimal postnatal environments in infancy and childhood, biological, psychological, and social-stressors in adolescence [6, 10, 11].

PTB individuals are also at an increased risk of exhibiting cognitive, neuromotor, or visual deficits later in life [8, 12, 13]. For instance, they have a substantial risk of developing cerebral visual impairment (CVI), a sensory deficit. In general, accurate vision and perception are paramount for navigating daily life, as the visual modality provides the richest information of all our senses [14]. In sighted individuals, it provides context to all the other senses and enables the construction of multi- or supramodal representations and predictions [14]. If this context is removed, it can elicit symptoms of psychosis, such as hallucinations, even in healthy individuals [15]. The visual sense also supports the development of motor abilities for navigating and interacting with our world [16]. In addition to cerebral visual problems, PTB children show poorer development of both fine and gross motor skills, which often persist into adulthood [17]. Poorer motor skills have been associated with higher levels of psychiatric symptoms and lower quality of life scores—underlining the importance of examining both visual and motor skills in relation to mental health [18].

Some studies have operationalized this as an IQ score falling more than two standard deviations below the age-group mean (e.g., ≤82 at age six) [9]. On average, children born preterm tend to score lower on standardized IQ tests than their term-born (FT) peers, with mean differences of approximately 12 points reported in certain studies [19]. In some cases, cognitive difficulties appear to persist into later childhood, with up to 50% of affected children still exhibiting such patterns at age 11 [9], and potentially extending into adulthood[19]. The importance of IQ scores is underlined by early scores measured in 2-year-olds being predictive of outcomes in adulthood [20]. These differences are not surprising, since most of the brain development occurs in the third trimester of pregnancy [21], which means that PTBs are born during or before this important and vulnerable period for brain development, which might give them a disadvantage from the starting line.

Although researchers and clinicians are aware of the outlined differences and factors related to PTB and specific birth circumstances, first studies report that the predictive accuracy of preterm events and factors favoring the development of a psychiatric disorder remains low [22, 23]. This low predictive accuracy might be attributed to relying on self-reports and examining already help-seeking individuals, which is a common limitation of the indicated prevention paradigm [5]. The focus on proximal predictors, such as functional decline and attenuated psychotic symptoms, may have led to an underrepresentation of distal risk factors like PTB [22].

Clear evidence for the role of PTB in psychiatric vulnerability comes from birth cohort studies [24]. A study examining adults born very premature (i.e., before 33 weeks’ gestation) found higher rates of psychiatric symptomatology, including increased positive, negative, cognitive, and behavioral symptoms, compared to FT-born adults [23]. These individuals were significantly more likely to fall into a high-risk psychopathology cluster, indicating a general psychiatric vulnerability rather than a disorder-specific risk [23]. These findings align with the neurodevelopmental model of psychiatric disorders, suggesting that altered brain connectivity in preterm individuals may contribute to their increased susceptibility to develop mental ill-health later in life [23].

Current approaches to early detection are predominantly informed by indicated prevention, with limited consideration given to the specific context of birth and psychometric assessments of low-threshold markers, such as visual and motor function. Indicated prevention may overlook low-threshold indicators, such as subtle early signs in visual and motor domains, which can sometimes be observed well before help-seeking behaviour begins. The integration of existing prevention paradigms, namely selective and indicated prevention, holds potential for enhancing early detection and overall health outcomes for individuals with PTB [5]. However, to date, a synthesis of potential concomitant developmental links between visual, motor, and cognitive skills after PTB and the risk of developing mental health problems in adolescence is lacking [23]. This review aims to address this gap by systematically examining the existing evidence for direct links between cognitive, visual, motor abilities and mental health outcomes.

## Methods

The present systematic review was conducted according to the Preferred Reporting Items for Systematic Reviews and Meta-Analysis (PRISMA) guidelines [25]. The protocol was registered in the International Prospective Register of Systematic Reviews (PROSPERO) database on 25.03.2024 (registration #513150).

### Literature search

A comprehensive literature search was conducted using multiple databases, including PubMed, Scopus, PsycINFO and the Cochrane Library, with a cut-off date of 6^th^ November 2023. Handsearching of reference lists of relevant articles and systematic reviews supplemented the systematic search (done by FS and LF). Filters ensured the inclusion of papers written in English. Our search included randomised and non-randomised trials, reviews, meta-analyses, and observational studies (cohort and case-control) meeting the inclusion criteria. We excluded animal studies, dissertations, books and book chapters, and opinion papers as well as articles without a representative sample size of preterm and/or low birth weight participants (N <25). Duplicate records were removed. Preterm birth was defined as birth before 37 completed weeks of gestation, in line with the World Health Organisation classification [1]. Globally, an estimated 15 million babies—more than 1 in 10—are born preterm each year [26, 27]. Preterm births are further categorised into extremely preterm (<28 weeks), very preterm (28–31 weeks), and moderate to late preterm births (32–36 weeks)[26].

### Study selection

The study selection process was managed using Covidence (www.covidence.org). At all stages reviewers (FS and LF) assessed articles for relevance to the research question and eligibility criteria. A two-step screening process was conducted, beginning with a title and abstract screening, followed by a full-text review to assess eligibility based on the predefined inclusion and exclusion criteria. The reviewers examined the texts to make final decisions based on predefined eligibility criteria. Disagreements between reviewers during this process were resolved by discussion amongst these two reviewers.

The search strategy was developed to capture studies linking mental health with birth-related factors such as preterm birth (PTB), as well as motor, visual, and cognitive abilities in adolescents. Inclusion and exclusion criteria were defined prior to data selection and extraction (Table 1). Articles were eligible if participants were born in 1980 or later, preterm (<37 weeks), had low birth weight (<2400 g), were at least 10 years old at the time of testing, and exhibited an IQ score >70. Studies involving participants with conditions like Down syndrome or cerebral palsy or with prenatal exposure to frequent illicit drug or alcohol use were excluded. We focused on original observational studies and excluded reviews, editorials, and small-sample studies (<20) to ensure methodological rigor.

**Table 1.** Inclusion and Exclusion Criteria for Study Selection

| Criterion | Inclusion | Exclusion |
| --- | --- | --- |
| Publication year | Participants born in January 1980 or later |  |
| Gestational age | Preterm (<37 weeks gestation) |  |
| Birth weight | Low birth weight (<2400 g) |  |
| Abilities assessed | Motor, visual and/or cognitive abilities tested |  |
| Age at testing | ≥10 years old at time of testing |  |
| Study types | Observational studies (cohort, case-control), randomized trials, perspective articles | Reviews, book chapters, editorials, opinion papers, small samples (<20) |
| IQ threshold |  | Intellectual disability (IQ<70) |
| Excluded conditions |  | Down syndrome, cerebral palsy, PVL, grade IV IVH |
| Excluded maternal factors |  | Maternal history of frequent illicit drug or alcohol use during pregnancy |

The initial literature search identified a total of 491 studies across all databases. Of these, we excluded 173 duplicates. The remaining 315 studies were screened by two independent reviewers (FS and LF) using title and abstract screening, which resulted in the exclusion of 202 studies. Subsequently, a total of 108 studies were subjected to full-text screening. We excluded 90 studies at this stage, as they did not meet all pre-registered inclusion criteria (Table 1).

This process resulted in a final sample of 18 articles analysed in this systematic review (Fig 1; Table 2).

**Fig 1.**
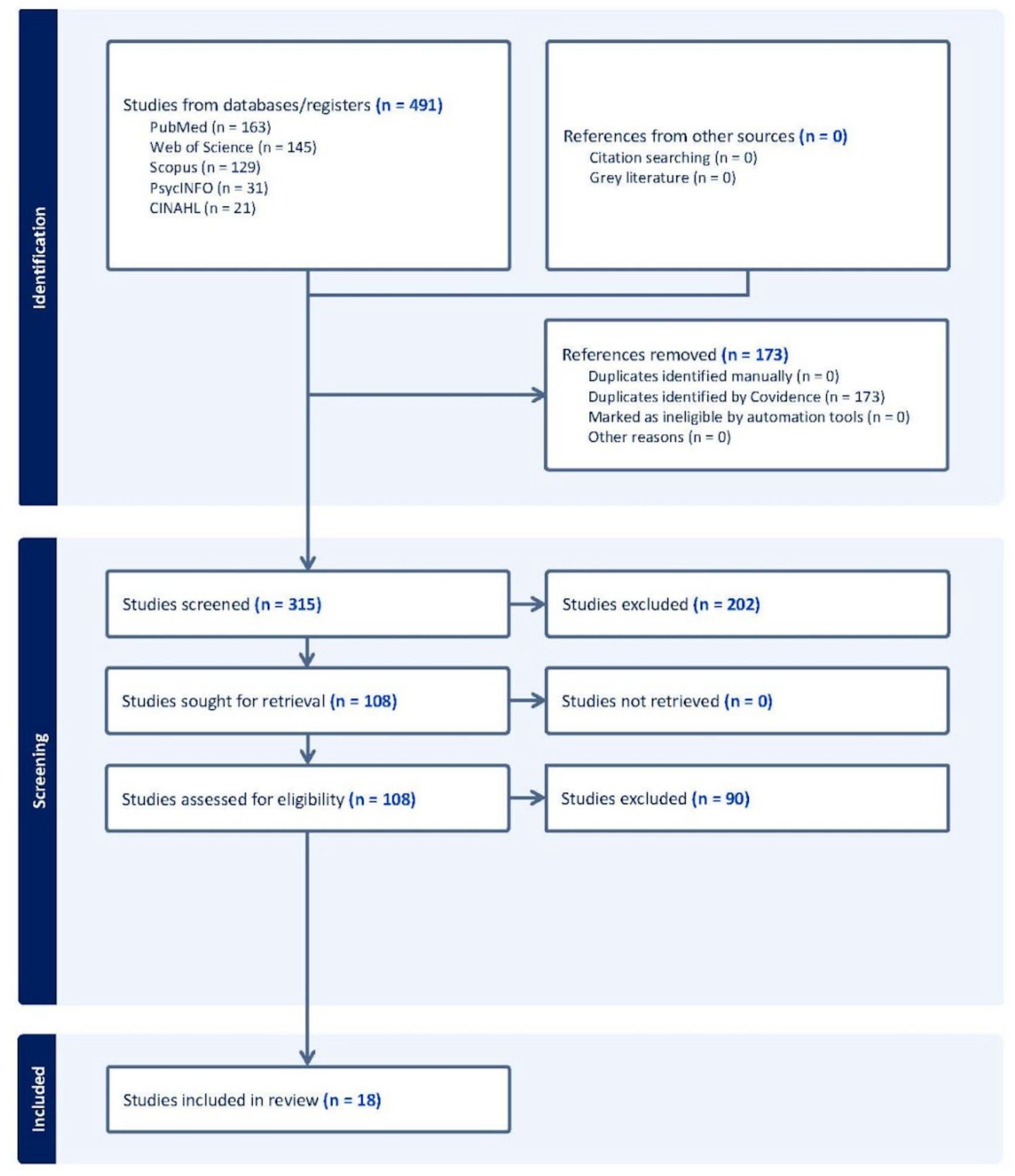
PRISMA-Flowchart of the study selection process. Flowchart of study selection based on PRISMA guidelines. Studies were excluded for the following reasons: sample too old (n=18), wrong form of publication (n=16), cohort sampling years not matching criteria (n=13), pediatric population (n=12), adult population (n=7), wrong patient population (n=7), adolescent data not available (n=4), missing key information (n=2), wrong outcomes (n=2), wrong language (n=1), and wrong study design (n=1).

**Table 2.**
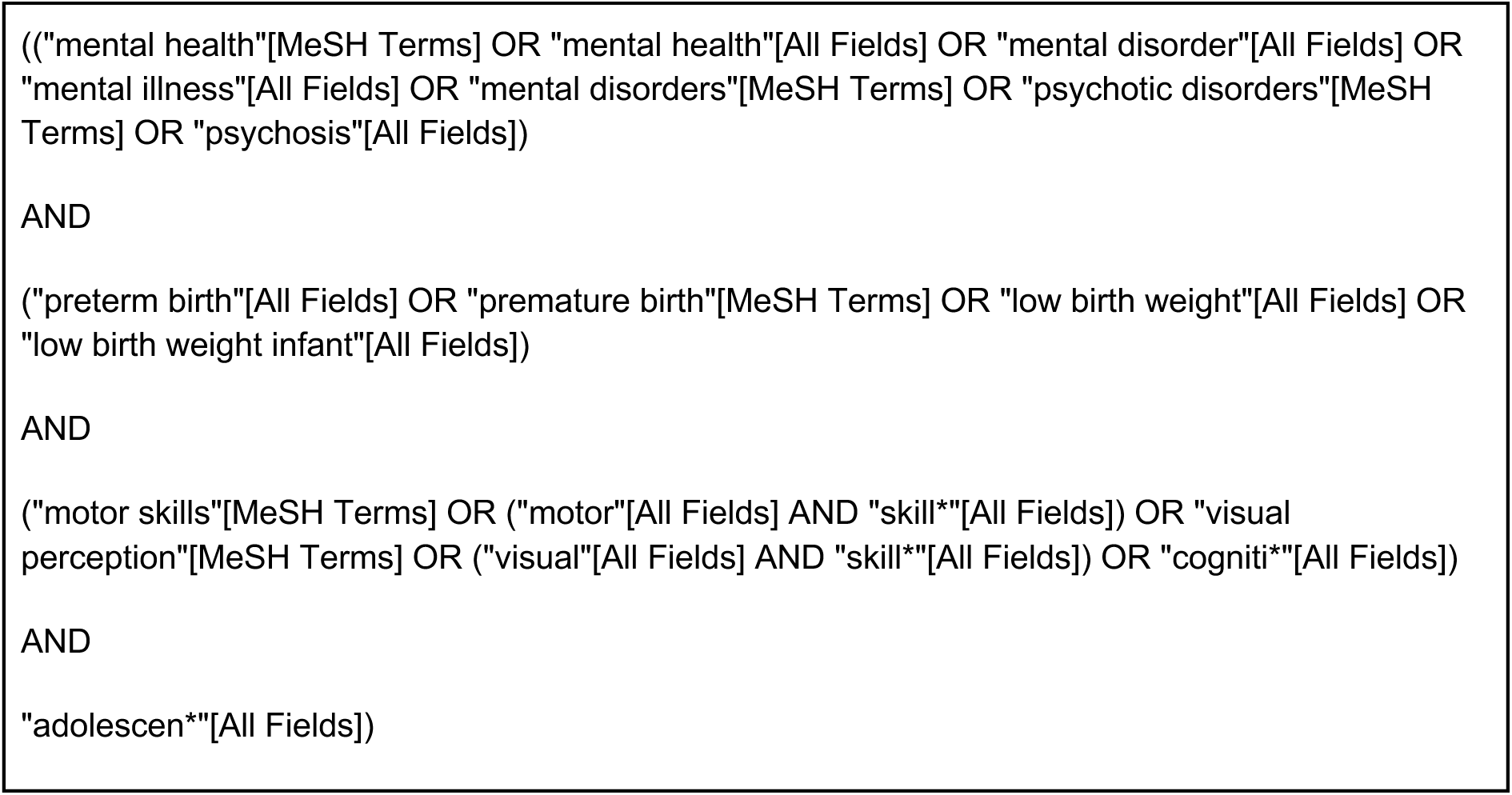
PubMED Example of Search String

**Table 3.**
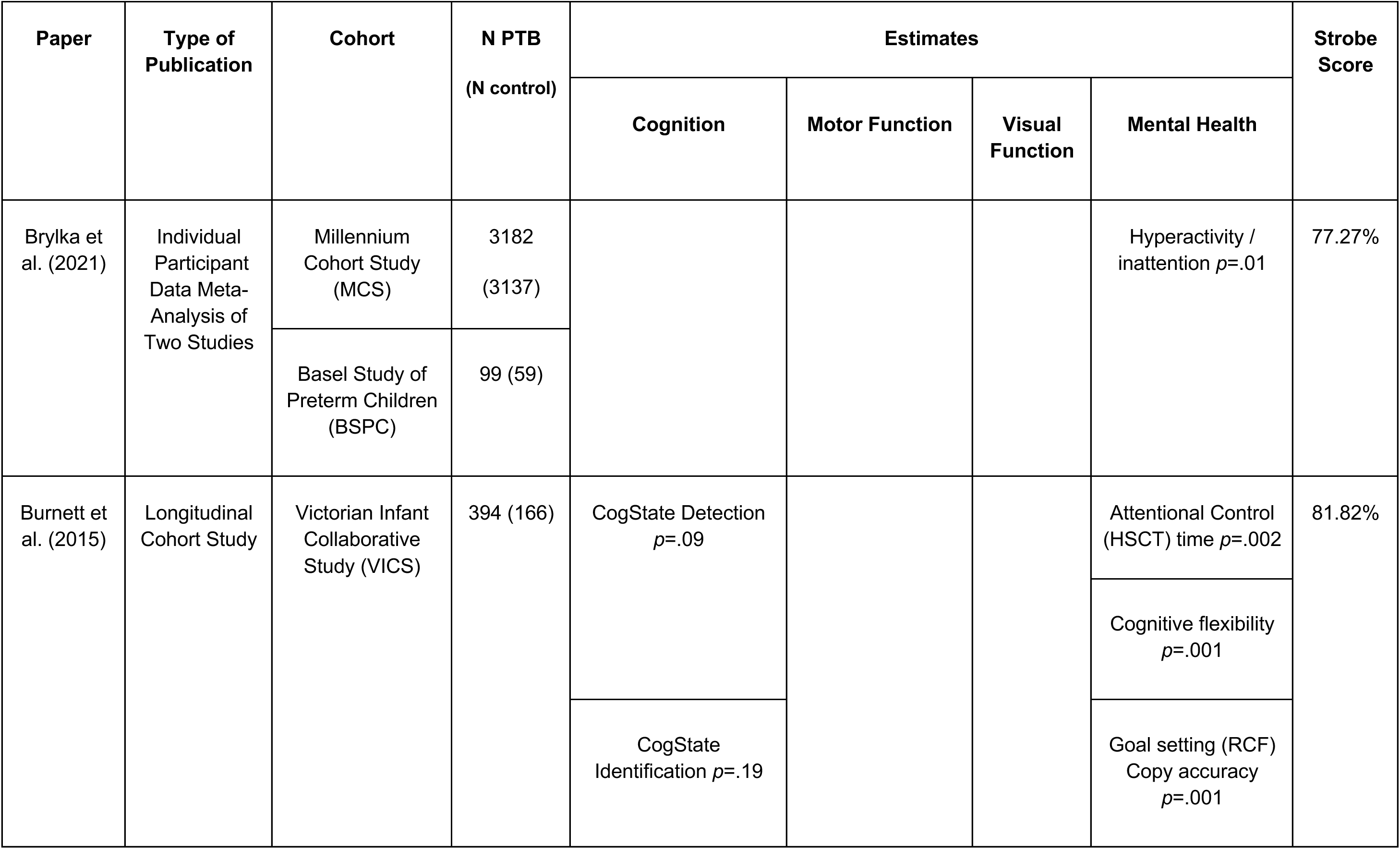
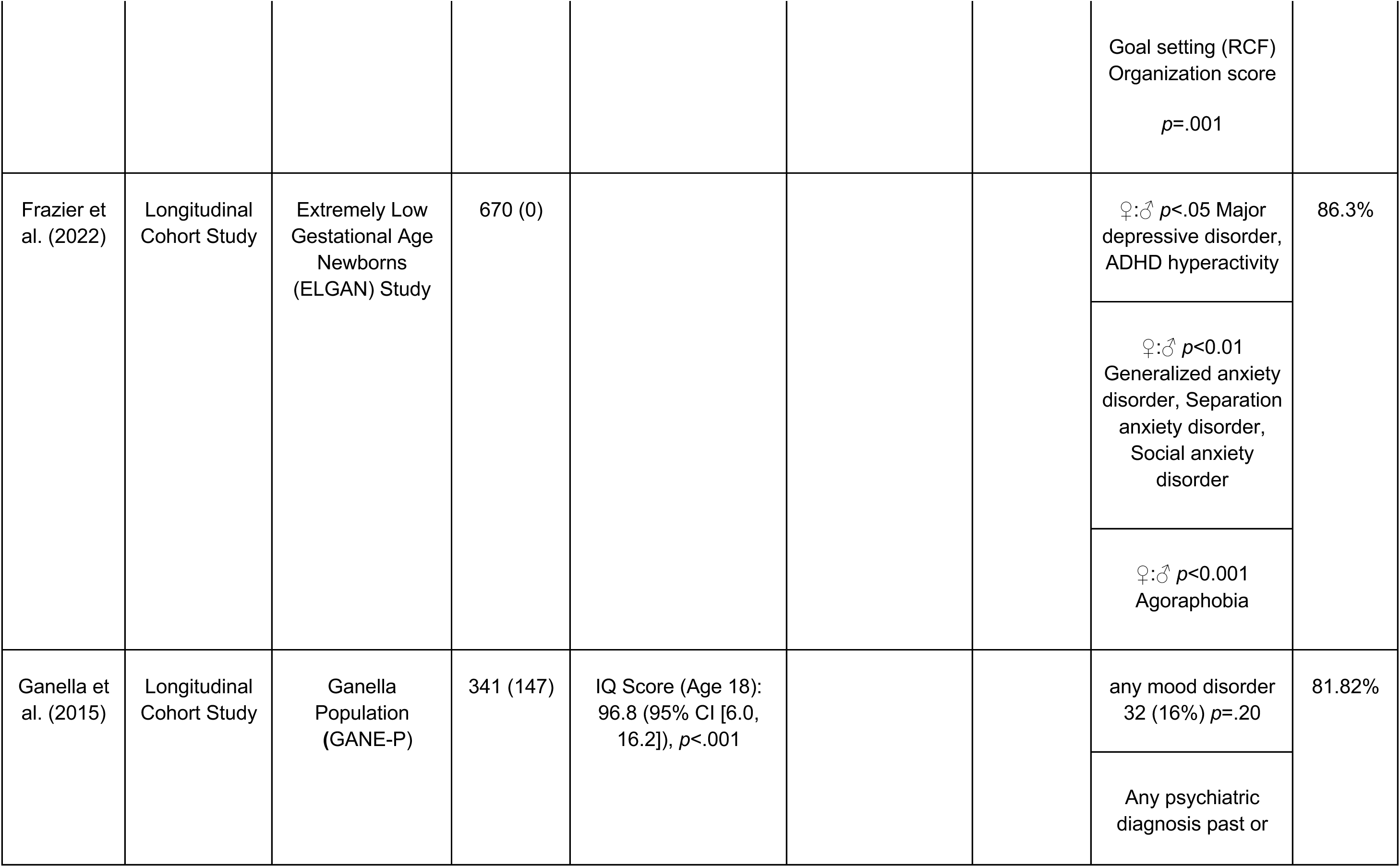
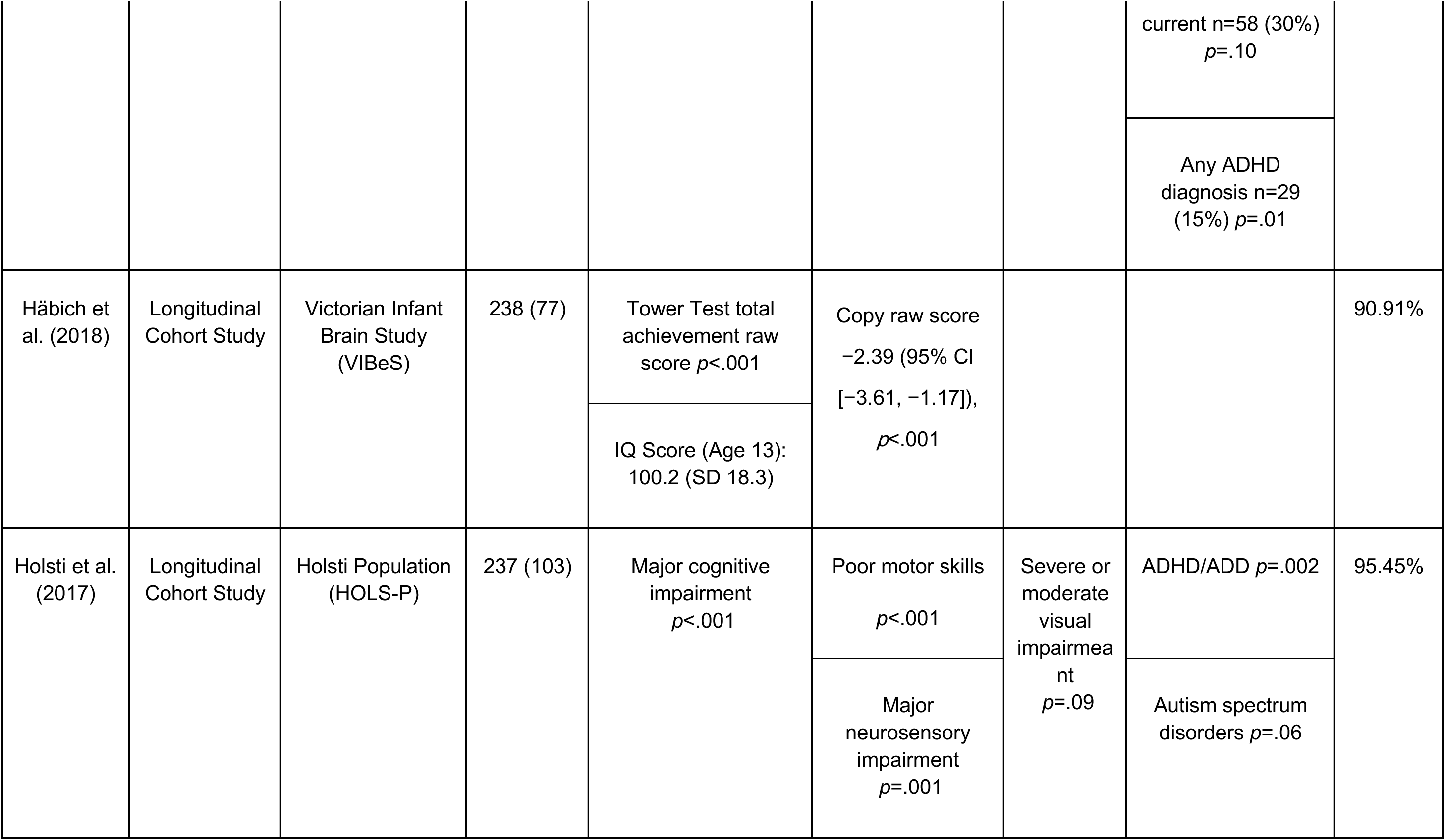
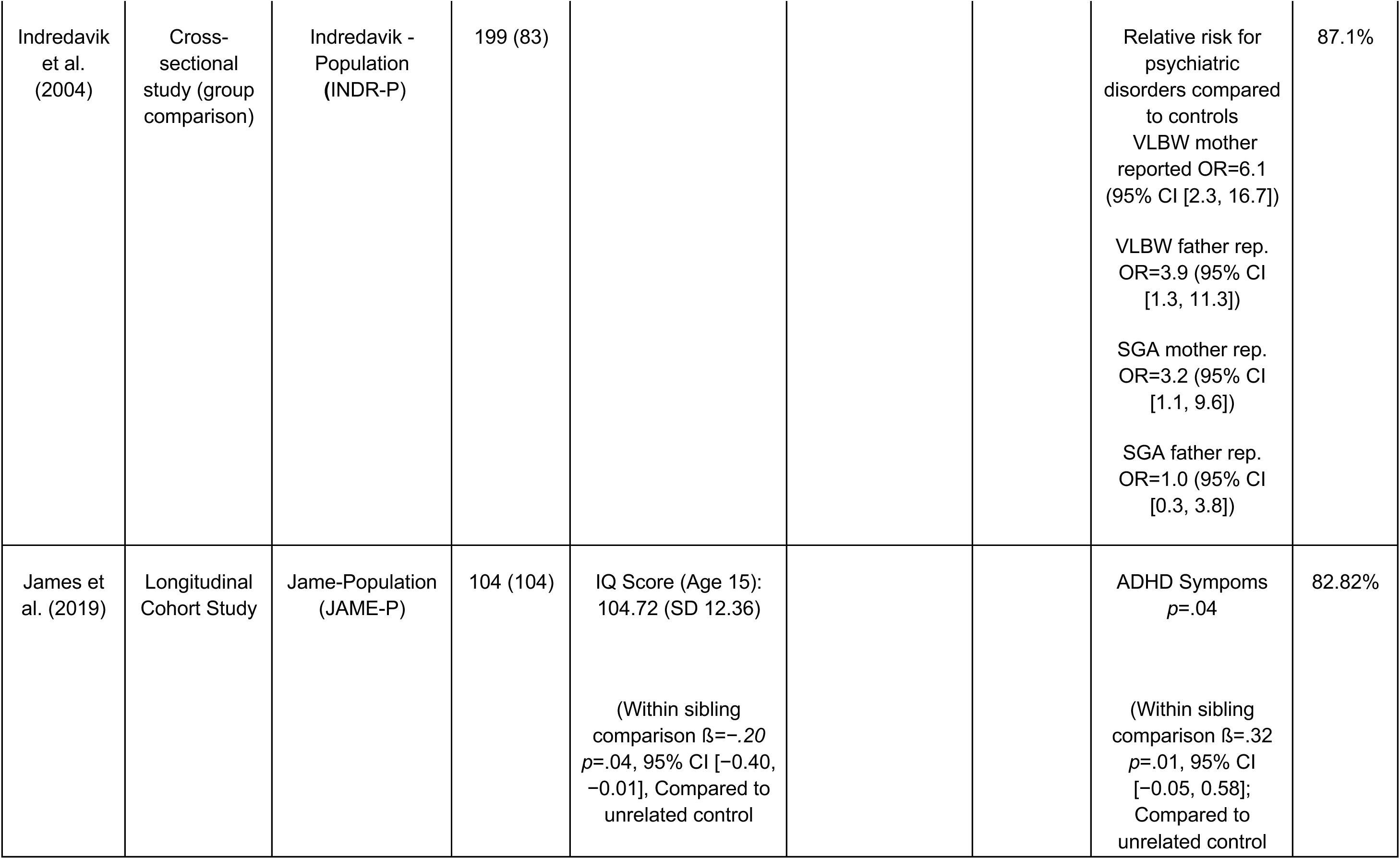
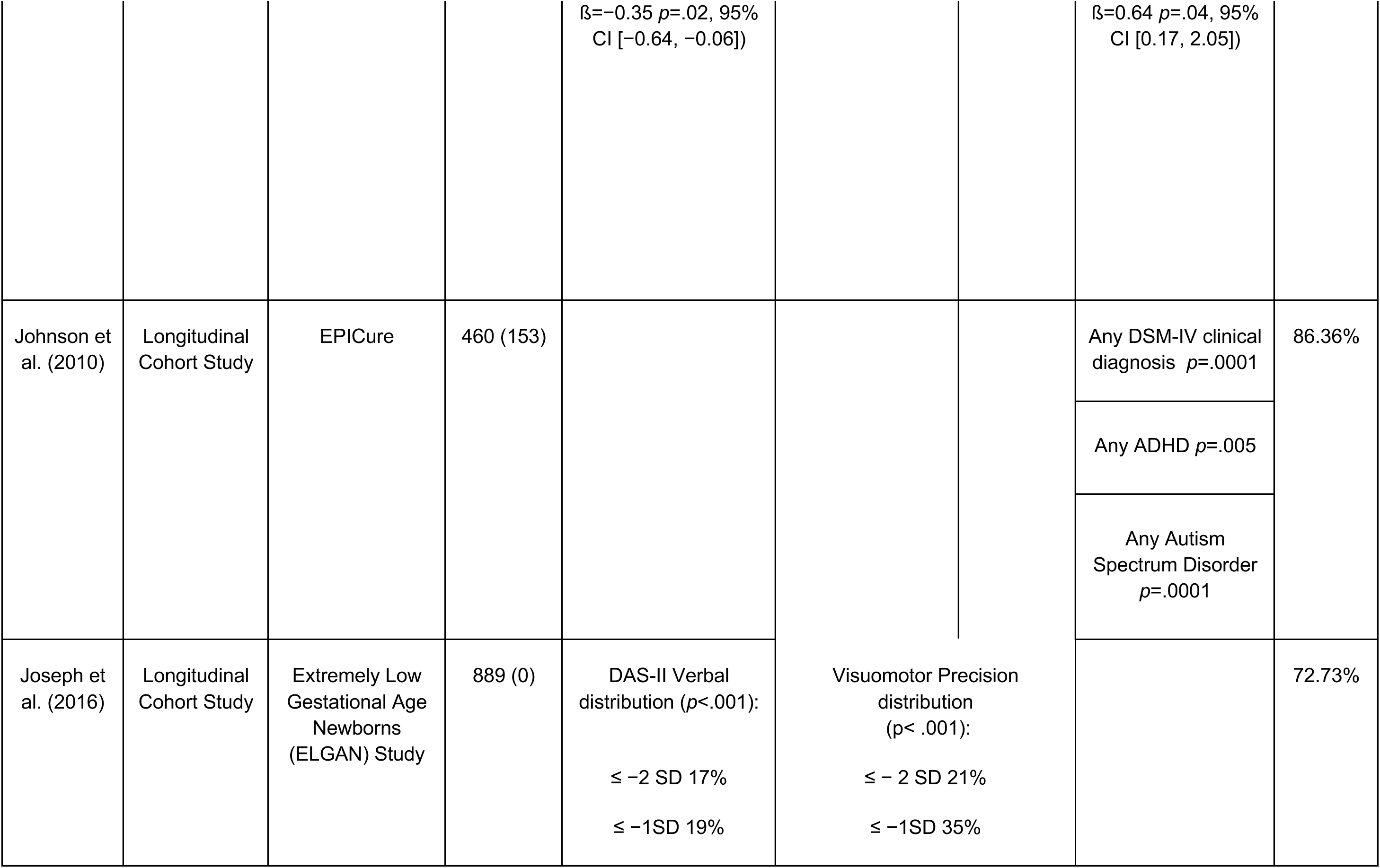
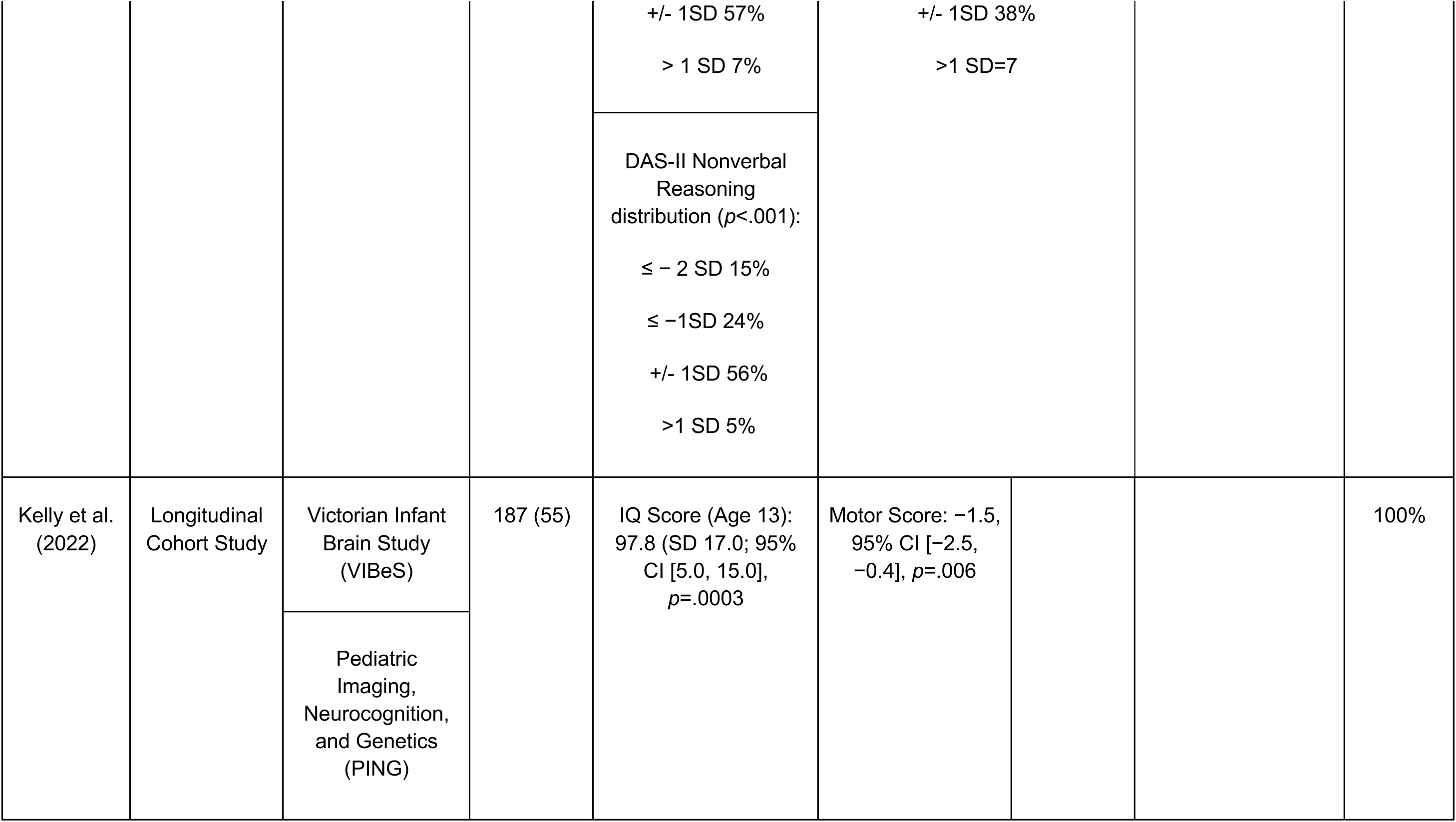
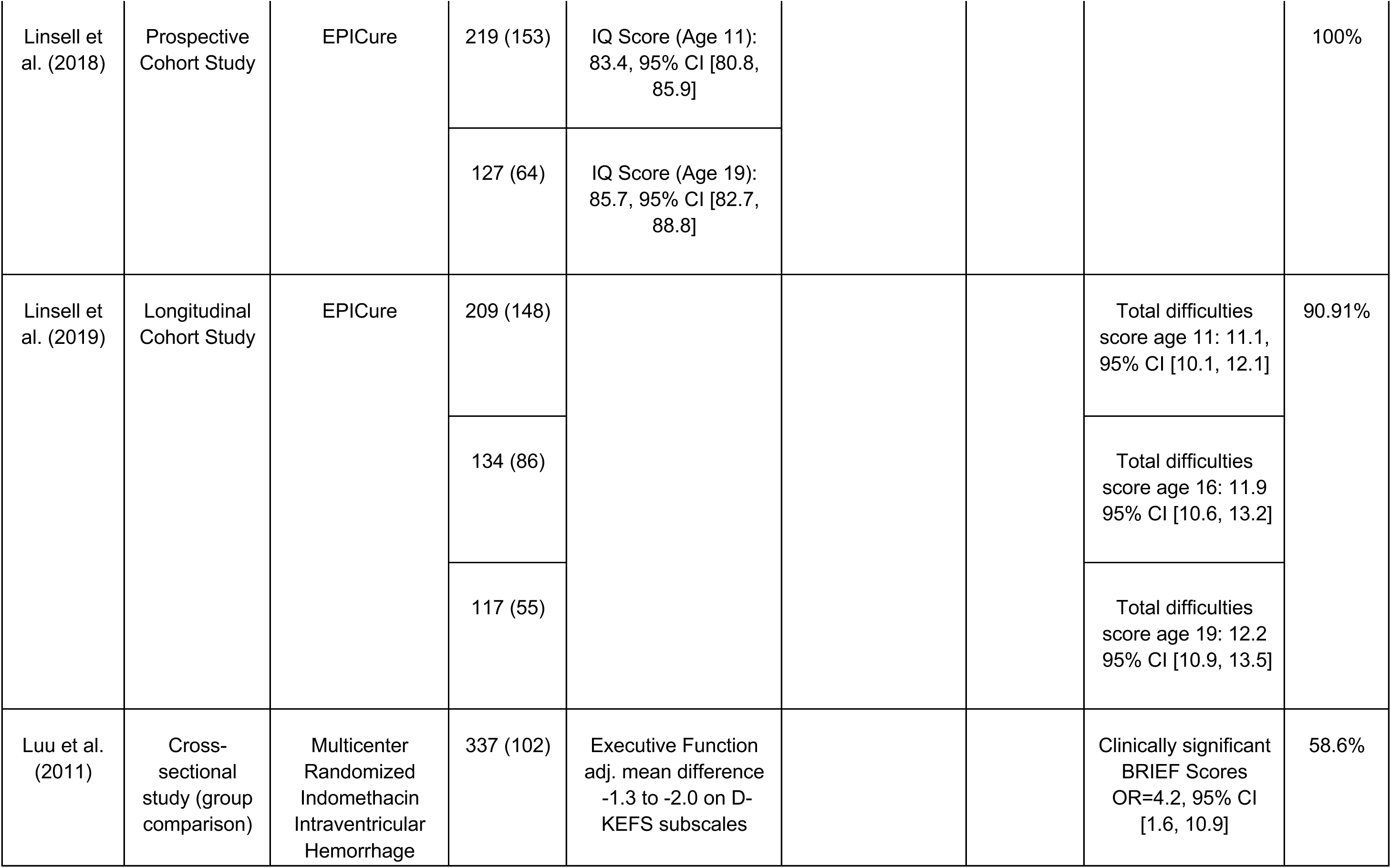
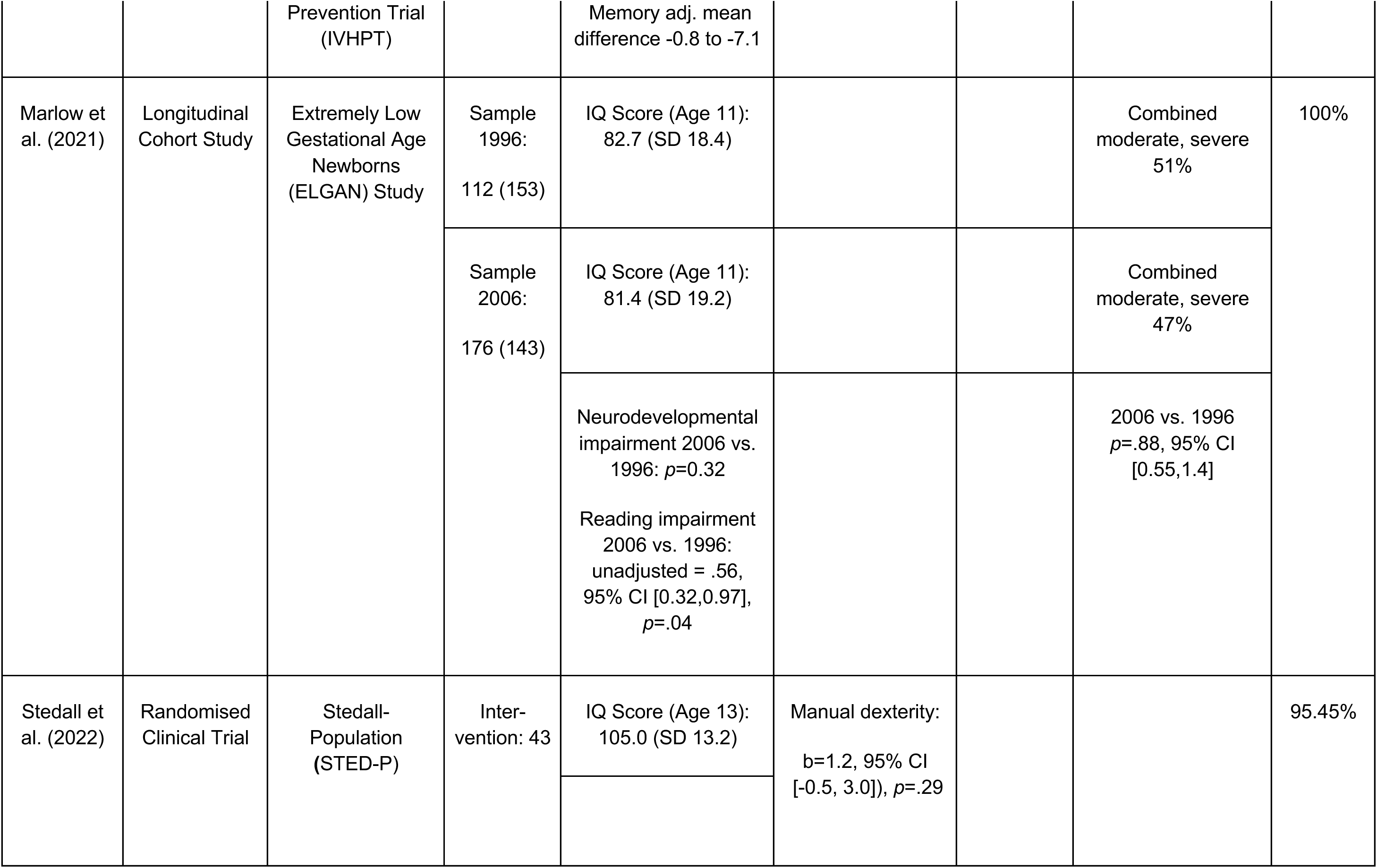
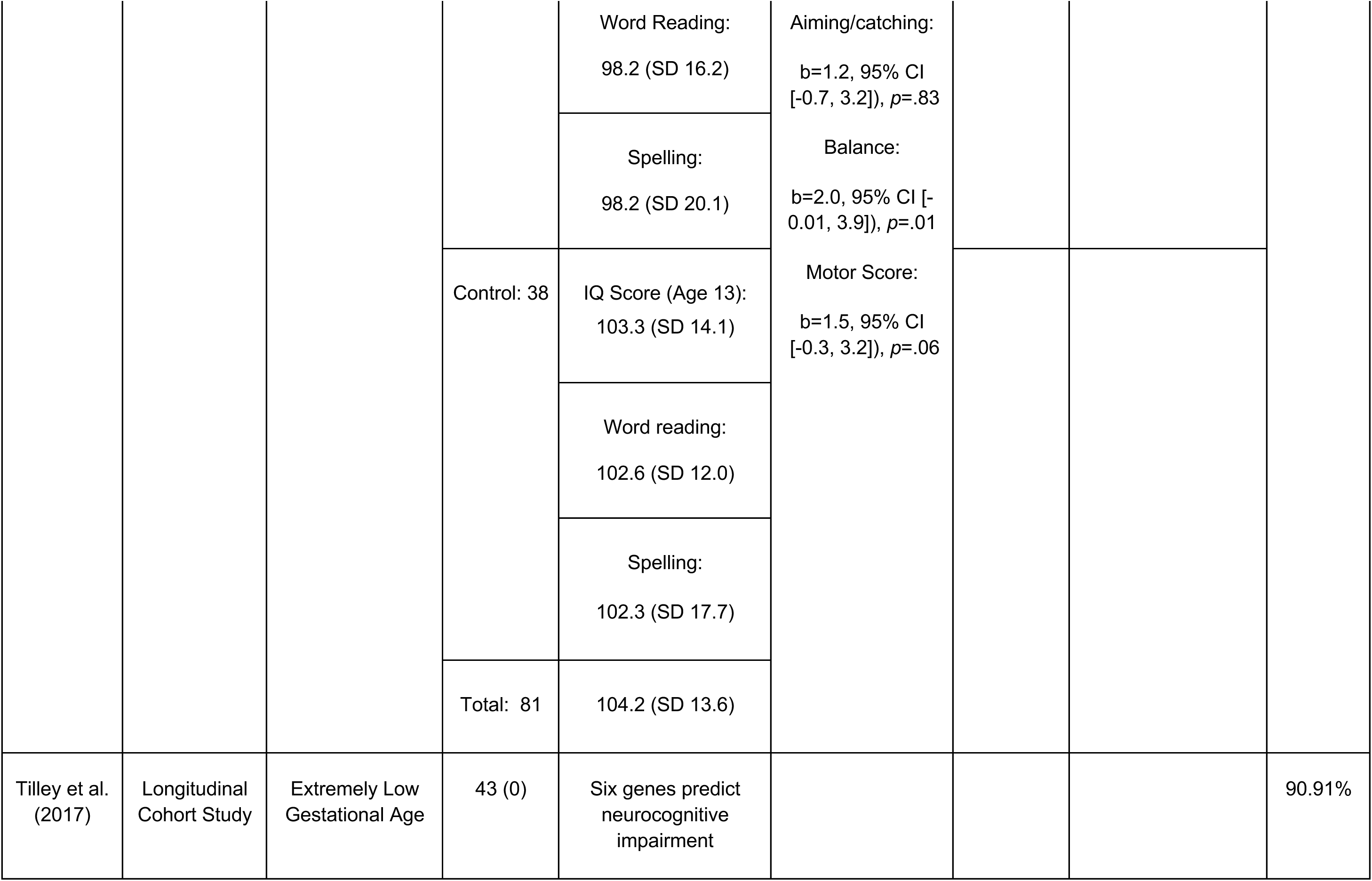
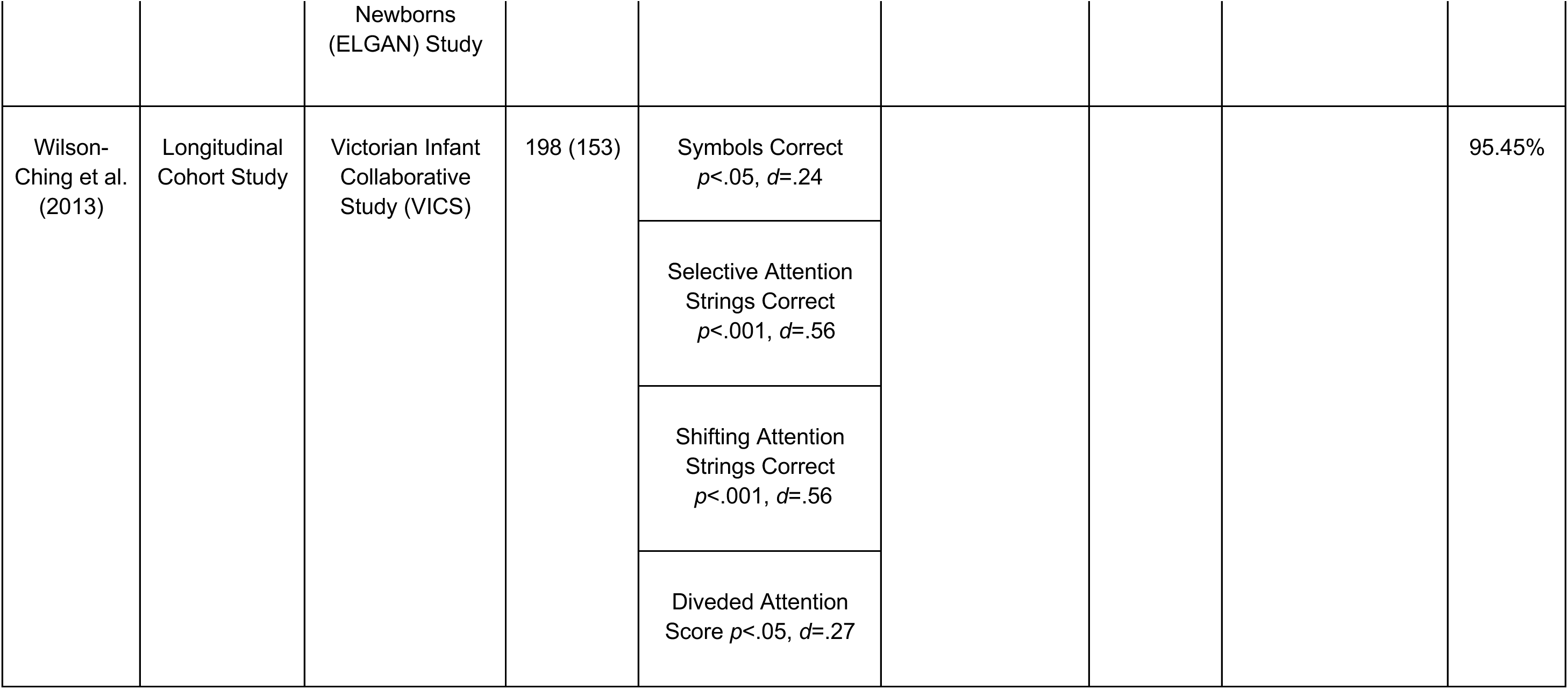
Table of findings

### Data extraction and quality assessment

The data extraction process involved the first author extracting the following variables for each included publication, if available: (1) name of the study; authors and publication year; (2) study design; (3) population; (4) analysed assessments or diagnostics; (5) findings. To quantify a study’s quality, we conducted the Strengthening the Reporting of Observational Studies in Epidemiology analyses (STROBE; [28]). Additionally, for each publication, we extracted all available data on cognitive, motor and/or visual abilities as well as mental health.

### Data synthesis

The data synthesis included all relevant data from all systematically identified and included studies. Given the wide variability in topics and data within the identified studies and domains, all reported findings are presented and compared in the context of the research question. Additionally, we conducted a meta-analysis of the IQ data available from the studies included in this systematic review.

### Meta analyses

The R packages meta, metafor, and ggplot2 were used for statistical analysis and visualisation. The figures illustrating the results of the meta-analyses (i.e., age-stratified comparisons of mean IQ scores and raincloud plot [29]) were created using RStudio (version 4.3.2) and MATLAB (version 2024b, The MathWorks Inc., Massachusetts, USA), respectively. Additional graphical adjustments were made using Adobe Illustrator.

Two publications, Stedall et al. [30] and James et al. [31], provided no IQ value for the control group. To fill these gaps in the meta-analysis, the missing values (i.e., means, standard deviations, sample sizes) were estimated by averaging across the available data for available control groups of the reviewed studies. These estimates of a “general” control group were then used as a proxy for the missing control groups in the studies [30, 31]. This imputation procedure is consistent with recommendations for handling missing data in meta-analyses when the number of missing values is small and the comparison groups can be assumed to be sufficiently homogeneous [32, 33]. The imputation was performed for the control group exclusively. Further, the study by Stedall et al. (2022) [30], comprised two groups of premature infants, one of whom received a physical early preventive care intervention. The two groups were tested concurrently and showed limited IQ differences (*M_Intervention_* = 105, SD = 13.2; *M_Non-intervention_* = 103.3, *SD* = 14.1). Since no significant difference in IQ score was observed for these two groups, we used the mean score across both PTB groups in this meta-analysis. This decision was made because both groups had similar conditions.

Together, the amended control data and PTB data formed the basis for two random effects models estimating 1) the standardised mean difference in IQ scores and 2) Hedges’ *g* as a measure of effect size. These random-effects meta-analyses were conducted using the restricted maximum likelihood (REML) estimator, based on k = 9 independent effect sizes. The meta-analyses were conducted on an aarch64-apple-darwin20 in RStudio (version 4.3.3; 2024-02-29, “Angel Food Cake”, [34]) using the metafor package version 4.8.0 [35]. The effect size model’s fit was slightly better (AIC_IQ_ = 60.62, BIC_IQ_ = 60.78; AIC*_g_* = 45.29, BIC*_g_* = 45.45). We report results from both models for transparency, as they may serve as a basis for future comparisons with alternative model specifications (e.g., fixed-effects models) and other studies reporting IQ scores.

## Results

The present systematic review identified 18 eligible studies published between 2004 and 2022 (Fig 2A). These studies included a total of 13.665 adolescents (PTB = 8.813, reported control = 4.852, three studies included no control participants [11, 36, 37]). Visual inspection shows that publication frequency has been increasing in recent years (Fig 2B). According to STROBE, almost all of the studies were of excellent or good quality, with only one study receiving fair scores (Fig 2C). These analysed data originate from several separate cohort studies, complemented by a few single original datasets (for an overview, see Table 1; Fig 2A). Some studies analysed data from separate cohorts within the same publication [38, 39].

**Fig 2.**
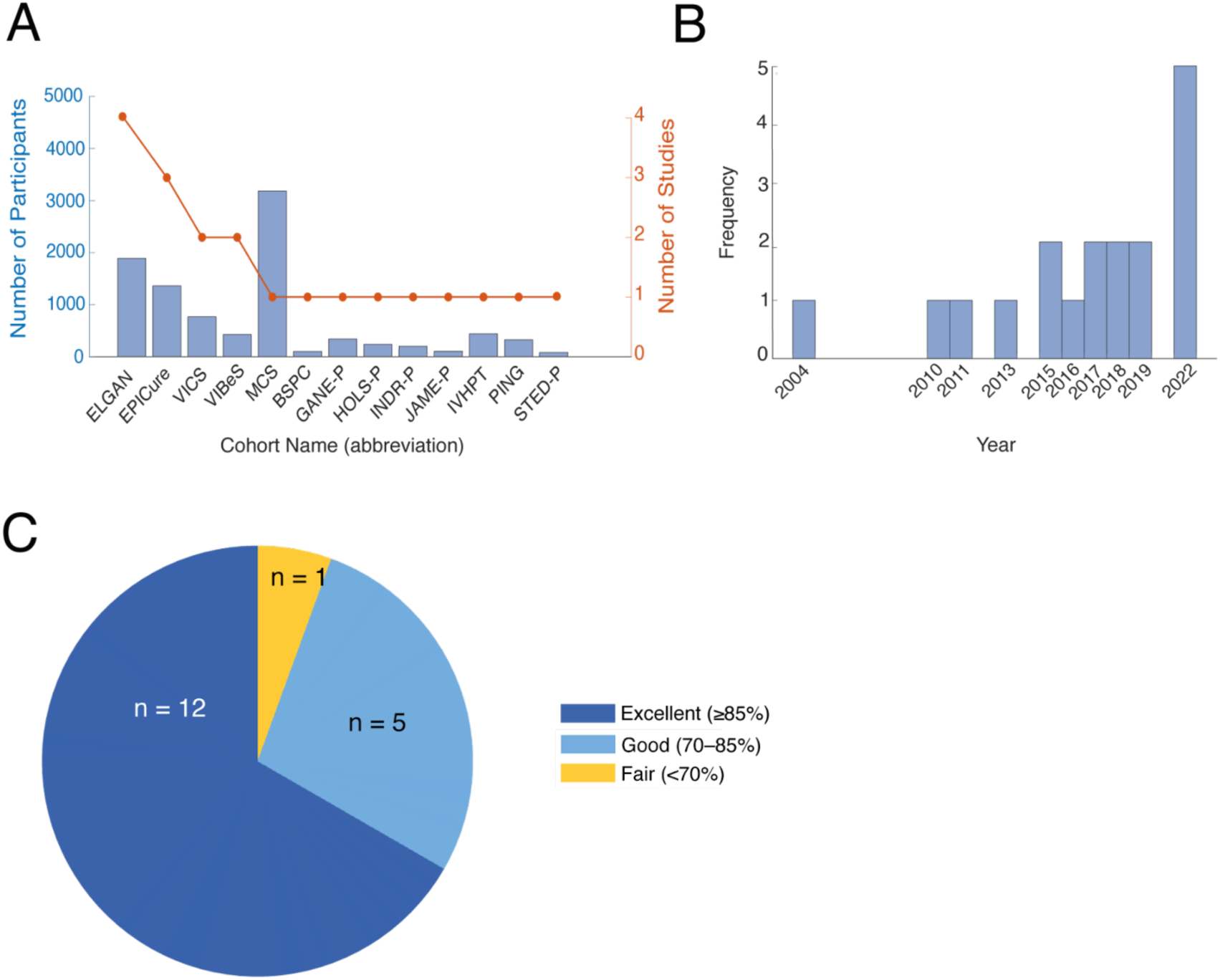
Number, temporal distribution, included participants and quality of included studies: **A)** Dual-axis plot displaying the number of participants and studies for each cohort, respectively (blue bars, left y-axis; red dotted line, right y-axis). The plot highlights differences between the cohorts’ sample size and the number of publications analysing their data. Bars represent the sample size per cohort. The red dotted line indicates the number of studies analysing each cohort’s data. **B**) Histogram showing the distribution of included studies by year of publication since 2004. The data indicate an increasing trend in the number of studies published over time with a very recent uptick in 2022. **C)** Pie chart illustrating the distribution of STROBE quality scores. (n = 12) rated as “Excellent” (≥85%), (n = 5) rated as “Good” (70– 85%), (n = 1) classified as “Fair” (<70%).

The included studies present a heterogeneous picture in terms of country of origin, participant age, and research team. The extremely low gestational age newborns (ELGAN) cohort appears in four included studies [9, 11, 11, 37]. The EPICure cohort is included in three studies [9, 40, 41]. In two cases, a cohort is reported in two included studies—one is the Victorian Infant Collaborative Study (VICS; [42, 43]) and the other being the Victorian Infant Brain Study (VIBeS; [39, 44]). Nine of the reviewed studies investigate a single cohort, or a cohort that is included in only one publication [30, 31, 38, 38, 39, 45–48].

### Cognition

Thirteen articles examined cognitive abilities of PTB adolescents [11, 30, 31, 36, 37, 39, 40, 42–49]. Holsti and colleagues [46] found major cognitive impairments to occur significantly more often in PTB adolescents (*p* < .001; [46]). Several studies examined IQ as a more detailed proxy of cognitive abilities.

Our meta-analysis of IQ raw scores revealed a mean IQ score of 93.47 points of PTB adolescents across studies, with a mean difference of -12.95 points between the groups (*k* = 9, *SE* = 2.77, 95% CI [–18.39, –7.51]; *p* < .0001; Fig 3A). An additional random-effects model of effect sizes corroborated this significant pooled reduction in IQ scores (*k* = 9, *g* = -2.28, *SE* = 1.06, 95% CI [-4.36, -0.30], *p* < .0317; Fig 3B). Between-study heterogeneity was substantial (τ² = 10.1111, *SE* = 5.08, τ = 3.18). Importantly, the I² value of 99.81% indicates that nearly all of the observed variance is due to true heterogeneity rather than sampling error. Cochran’s Q-test for heterogeneity was statistically significant (Q(8) = 912.74, *p* < .0001), providing more evidence for differences in reported IQ outcomes among preterm populations reflecting meaningful between-study differences rather than random variation. Interestingly, the effect size of two PTB groups reported within the same study was substantially larger than the remaining ones ([41]; Fig 3B). Although this study increases the observed overall effect, a clear reduction in PTB adolescents would be evident still, as only two studies report a non-significant reduction (Fig 3A, C).

**Fig 3.**
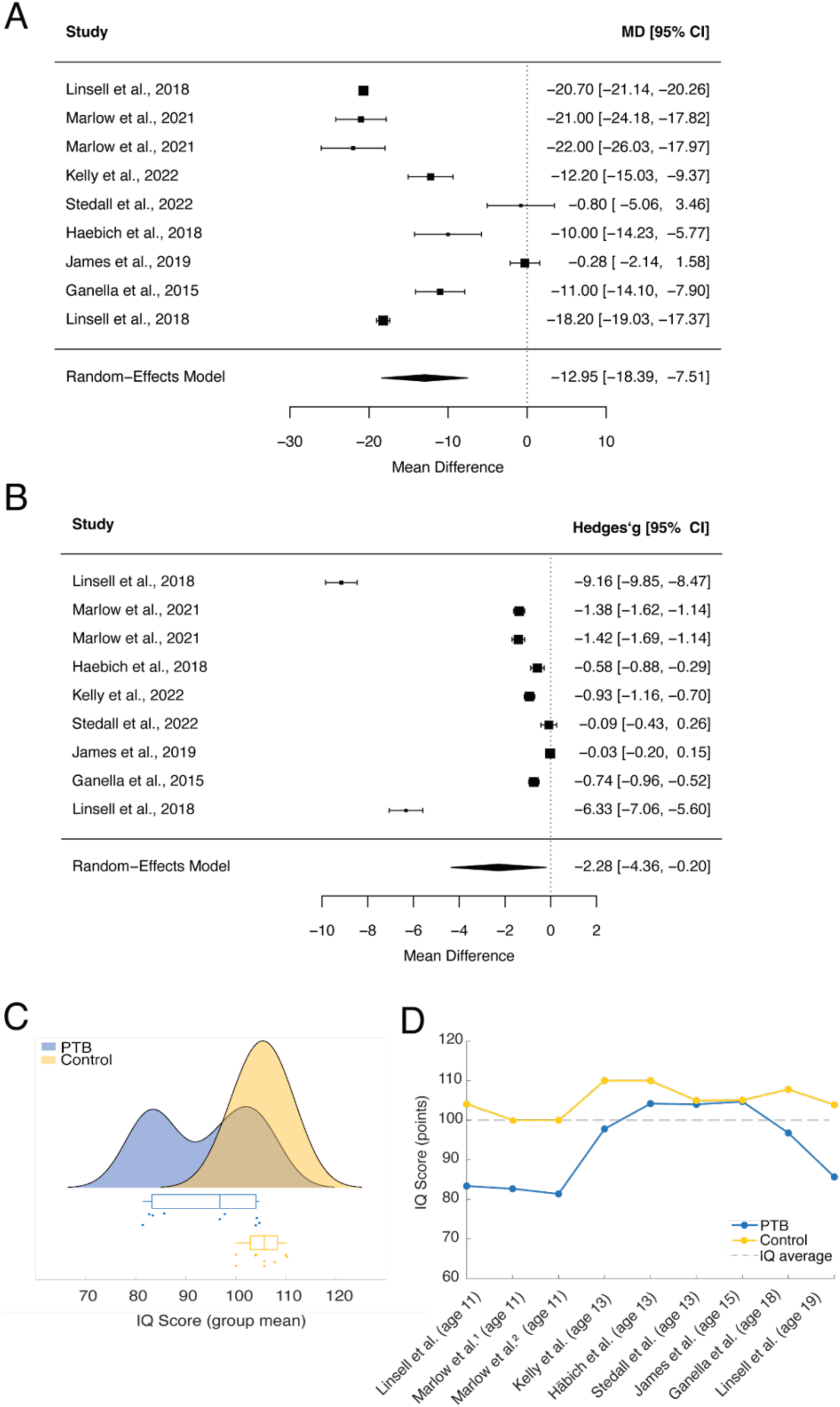
Meta-analyses of mean differences (MD) in cognitive IQ test scores (points) and Hedges’ *g* across reviewed studies. **A)** Forest plot illustrating the MD in IQ test scores with their corresponding 95% confidence intervals for each examined study. Sorted by ascending age. Pooled effect sizes based on a random-effects model (*k* = 9; REML estimator). The overall effect was statistically significant (*g* = -12.95, *SE* = 2.77, 95% CI [-18.39, -7.51], *p* < .0001). Between-study heterogeneity was substantial (*τ²* = 66.89; *I²* = 99.12%; Q(8) = 588.73, *p* < .0001), indicating considerable variation in effect sizes across studies. Model fit indices: AIC = 60.62, BIC = 60.78. **B)** Forest plot illustrating Hedges’ *g* across reviewed studies with their corresponding 95% confidence intervals for each examined study. This list is sorted by ascending age. Pooled effect sizes based on a random-effects model (*k* = 9; REML estimator). Model fit indices: AIC = 46.97 and BIC = 47.13 **C)** Raincloud plot [29] of sample mean IQ scores depicted in panel A, and boxplots comparing scores for PTB (blue) and FT samples (yellow) independent of age. **D)** Comparison of mean IQ scores between PTB (blue) and FT control groups (yellow) across the included studies. Where no FT data was available [13], we used the mean of the available control groups as a comparator.

Further analyses of the temporal evolution of IQ differences throughout adolescence illustrated that IQ score differences follow a U-shape, with PTB individuals exhibiting larger reductions at age 11 (Fig 3D, Δ = -18.87 points, *M*_PTB_ = 82.7, 95% CI [23.13, -14.60], [13, 40]), and 19 (Δ = -18.2 points, *M*_PTB_ = 85.7, 95% CI [82.7, 88.8]; [40]). IQ differences between PTB and their FT peers decrease towards mid-adolescence at age 13 (*Δ* = -9.97 points, *M*_PTB_ = 101, 95% CI [91.88,110.12]; [30, 39, 44]) and 15 (Δ = -4.33 points, *M*_PTB_ = 104.72, 95% CI [-0.64, -0.06]; [50]).

Several studies also examined specific cognitive abilities, such as executive functions and attention. PTB adolescents exhibited intact planning skills at age 13 with a score of 9.9 out of 10 on the Delis-Kaplan Executive Function System (D-KEFS; [51]). On the contrary, adjusted mean differences on executive function tasks across all D-KEFS subscales indicated that impairments also exist in the PTB group, albeit at varying levels. On the one hand, the least impairment was observed in the Category Fluency subtest (*M* = 9.7, *SD* = 3.7), with 6.3% of children scoring two standard deviations below the norm (*aMD* = -1.3, 95% CI [-2.1, -0.5]). The most significant impairment was observed on the Color-Word Inhibition subtest (*M* = 7.9, *SD* = 3.8), where 17.6% of children scored two standard deviations below the norm (*aMD* = - 2.0, 95% CI [-2.8, -1.2]; [48]). As well, PTB adolescents exhibited significant planning and organisation difficulties on the Behaviour Rating Inventory of Executive Function (BRIEF; *β* = 2.5, 95% CI [1.2, 5.1]; [42, 52]). More cognitive performance measures showed significant impairments in selective, shifting, and divided attention (*p* < .001, *p* < .001, *p* < .05, respectively; [43]). Neurocognitive impairment could even be predicted by six genes, as observed in a study involving preterm-born individuals [37].

Taken together, these studies highlight cognitive challenges in PTB adolescents, particularly in general IQ, executive function, and attention [44, 53]. These are particularly prominent in the beginning and end of adolescence, and may persist in adulthood.

### Motor function

Motor functioning relies on integrating neurosensory input to interpret environmental cues and generate appropriate responses. Five included studies examined motor function with a sample size of more than 20 participants [30, 36, 39, 44, 46].

Two studies found a considerable prevalence of general motor impairment in adolescence [39, 46]. Specifically, during earlier adolescence (i.e., age 10-15), one study showed a generally higher prevalence of poor motor skills in PTB adolescents without neurosensory difficulties using the M-ABC [46]. Also at age 10, the majority of PTB adolescents showed performance considerably below average on the Visuomotor Precision task (21% of participants scored ≤ (−2) SD, 35%: > (−2) and ≤ (−1) SD, 38%: > (−1) to ≤1 SD, and 7%: >1 SD; [36]). At age 14, another study reported a significant deficit in balance (*β* = 2, 95% CI [−0.01, 3.9], *p* = .01; [30]). However, this study’s total standard score for balance was non-significantly reduced, requiring cautious interpretation (*M* = -1.5, 95% CI [0.3, 3.2], *p* = .06; [30]).

Holsti et al. [46] point out that a significant proportion of PTB adolescents have functional limitations like neurosensory impairments (NSI; *aOR* = 15, 95% CI = [6.1, 37.2], *p* < .001; [46]). NSI, defined as moderate or disabling cerebral palsy, blindness, moderate to severe visual impairment, or auditory impairment requiring bilateral hearing aids or a cochlear implant, were also reported to be unchanged in PTB adolescents 1996 or 2006 (*p* = .32; [13]). Specifically, fine motor skills, indexed by the Rey Complex Figure Test (RCFT; [54]) showed a significant reduction in the raw copy score at age 13 (*M*_PTB_ = 24.94, *SD* = 4.93, *M*_FT_ = 27.28, *SD* = 3.80; Δ = -2.39, 95% CI [-3.61, -1.17], *p* < .001; [42, 44]). As well at age 13, PTB adolescents demonstrated significantly poorer motor skills compared to their full-term peers (*M*_PTB_ = 9, *SD* = 3, *M*_FT_ = 10, *SD* = 2; Δ = -1.5, 95% CI [-2.5, -0.4], *p* = .006; [39]).

As a proxy of general motor issues, PTB adolescents demonstrated consistently lower levels of physical activity, reporting an average of 10 minutes less activity per day than their FT counterparts [38]. More specifically, a group of PTB adolescents with NSI was significantly more likely to receive physical/occupational or other therapy compared to their PTB counterparts without NSI (*aOR*_with_NSI_ = 22.1, 95% CI = [2.9, 166.8], *p* < .001; *aOR*_without_NSI_ = 11.1, 95% CI = [1.4, 87.7], *p* = .02; [46]). This latter finding suggests physical limitations severe enough to require therapy.

Contrarily, a higher likelihood of NSI was neither found in the Basel Study of Preterm Children (*p* = .563; [55]) nor the Millennium Cohort Study (*p* = .197; [55]). It is noteworthy that the latter two cohorts comprise more FT births than expected in the population, which may have reduced statistical power to detect smaller differences. Efforts counteracting motor issues can be successful in this population, as positive effects of interventions illustrate. Stedall et al. [30] showed that motor outcomes including manual dexterity (*M*_manual_dexterity_ = 7.5, *SD* = 3.4; *aMD* = 1.2, 95% CI [-0.5 to 3.0], *p* = .29) and balance (*M*_balance_ = 9.1, *SD* = 3.6; *aMD* = 2.0, 95% CI [0.01 to 3.9], *p* = .01) could be improved in PTB adolescents following a physical activity intervention.

In sum, the analysed studies provide evidence for an increased prevalence of motor deficits and less physical activity in PTB adolescents. Though, the evidence for an increased risk of neurosensory impairments is not unequivocal.

### Visual function

Three of the included studies investigated the effects of PTB on functional visual abilities including visual perception, visuo-motor control and visuo-spatial planning in adolescence [36, 44, 46]. They demonstrate that PTB adolescents have an increased risk of scoring between one and two standard deviations below the FT population mean on most visual tests [36]. Developing proficient visual abilities, such as visual perception and mental flexibility, are particularly challenging for PTB adolescents [36]. One study observed a non-significant trend (*p* = .09) toward moderate to severe visual impairment in some children, with a few exhibiting visual acuity below 20/200 in their better eye without correction [46]. Lastly, PTB adolescents were less likely to improve their conceptual organisation score, even after adjusting for baseline group differences [36]. A gender difference was reported for visuo-spatial organisation skills, where especially PTB males showed inferior performance [44].

The evidence, albeit preliminary due to the small number of studies, suggests that PTB adolescents may be at increased risk of suffering from moderate to severe visual impairment. Specifically, functional visual abilities may be impacted, as evidenced by lower performance in visual perception and visuo-spatial organisation compared to their age-matched FT peers.

### Mental ill-health

Eleven studies have investigated neurodevelopmental disorders, cognitive impairment and mental health in PTB adolescents [9, 11, 38, 40–42, 45–48, 50, 56]. These studies cover mild to severe mental health issues.

PTB adolescents exhibit a higher prevalence of developing psychiatric disorders suggesting increased emotional and psychological vulnerability. Frazier et al. [11] reported that 15% of PTB adolescents had received a diagnosis of one psychiatric disorder, 9% had been diagnosed with two co-occurring disorders, and 8% with three or more diagnoses, resulting in a total of 33% of PTB adolescents having been diagnosed with at least one psychiatric disorder. This prevalence exceeds global estimates for psychiatric disorders in the general population of children and adolescents considerably, which range between 10–20% [57]. The severity of symptoms requiring treatment is underlined by PTB adolescents presenting with psychiatric or neurodevelopmental symptoms being significantly more likely to engage in regular visits with counsellors, psychologists, or social workers compared to those without such symptoms (*p* = .04, *p* = .08; [11]). Furthermore, significant sex differences were reported, with girls being more frequently diagnosed with major depressive disorder (6% vs. 2%; *OR* = 0.3, *p* < .05), generalised anxiety disorder (11% vs. 5%; *OR* = 0.4, *p* < .01), agoraphobia (7% vs. 1%; *OR* = 0.2, *p* < .001), separation anxiety disorder (6% vs. 2%; *OR* = 0.3, *p* < .01), and social anxiety disorder (8% vs. 3%; *OR* = 0.3, *p* < .01), but less often with the hyperactive subtype of ADHD (1% vs. 4%; *OR* = 4.2, 95% CI [1.2, 15.0]; [11]).

Attention deficits and hyperactivity deserves specific attention within PTB adolescents, with ADHD being particularly prevalent (7-23%) and clinically relevant [11]. Parent and teacher reports support more frequent ADHD symptoms in PTB compared to FT peers (PTB = 25%; [47]). In addition, at age 15, 18% of the PTB adolescents met criteria for ADHD, with a significant sex difference in the combined subtype (*p* = .004) and the hyperactive subtype being more common in boys than girls (*p* < .001). Among those diagnosed with Autism spectrum disorders at age 10, 46.2% also met criteria for ADHD, and 61.5% of girls with Autism spectrum disorder had comorbid anxiety disorders—more than double the rate of girls without ASD (26.7%; [11]).

Comparing PTB and family vulnerability hypotheses of attention deficits, we find an attenuated prevalence of ADHD in siblings (*β* = 0.32, *p* = .01, 95% CI [0.05, 0.58]) compared to unrelated individuals (*β* = 0.64, *p* = .04, 95% CI [0.17, 2.05]; [31]). This result suggests that shared genetic or familial factors may not account for the observed comorbidity. Contrary, PTB individuals were more likely to receive an ADHD diagnosis than their siblings (*p* = .01), providing evidence for a specific vulnerability due to PTB [31].

While attention deficits and hyperactivity are prevalent in PTB adolescents, engaging in physical activity can play a protective role in their mental health. Specifically, increased physical activity was linked to greater psychological well-being (*β* = 0.05, 95% CI [0.019, 0.087; [38]) and more favourable self-perception (*β* = 0.06, 95% CI [0.028, 0.094]; [38]).

The findings emphasise the considerable impact of psychiatric and neurodevelopmental disorders on adolescents born preterm, including elevated rates of psychiatric diagnoses and pronounced gender disparities. Of particular note is the high prevalence of attention deficits and hyperactivity in this group, which frequently co-occurs with other neurodevelopmental conditions. This overlap complicates the isolation of specific developmental pathways and constitutes a potential confounding factor in studies aiming to assess the long-term impact of prematurity on cognitive and psychiatric outcomes.

## Discussion

The present systematic review underlines that individuals born preterm exhibit lower performance on cognitive and motor tests, and that these deficits remain evident into adulthood. Our findings also reveal an association between PTB and psychiatric disorders, particularly regarding ADHD, anxiety, and autism [9, 11, 38, 46, 50]. These findings are corroborated by recent WHO reports linking PTB to an increased risk of developing psychiatric disorders including ADHD, anxiety, and affective disorders [8, 9, 50]. Psychiatric disorders can also hinder academic performance and cognitive functioning of PTB adolescents, suggesting the inclusion of cognitive evaluations in early detection approaches as early as adolescence [50].

The reviewed studies show persistent executive function impairments in PTB adolescents, including deficits in working memory, planning, and cognitive flexibility. These cognitive difficulties are evident in standardised testing and parent-reported assessments, and can significantly impact everyday functioning [44, 48]. The presence of these impairments is indicative of broader challenges, with associated symptoms including attentional difficulties and a decline in academic performance [30]. Longitudinal data from birth until late adolescence suggest that cognitive deficits persist despite advancements in neonatal care, indicating limited compensatory neuroplasticity [49]. Neuroimaging studies in adults born preterm reveal structural brain differences, particularly reduced white and grey matter volumes in regions like the thalamus and striatum. These alterations may result from disruptions in early brain development, which can persist and become more noticeable in adulthood. These volume reductions are associated with cognitive impairments in memory, processing speed, and executive function. Such alterations reflect long-term disruptions in brain development rather than short-term effects of preterm birth [44, 53]. The latter findings emphasise the enduring impact of PTB on neurocognitive long-term outcomes and highlight the need for continued evaluation and support long after neonatal care has “ended”. Whether these persistent differences in cognition and executive functions constitute an increased vulnerability to develop mental ill-health remains to be seen.

The general cognitive challenges we identified in this review hint at an increased psychological vulnerability, as they indirectly support the known relation between lower cognitive performance and elevated psychopathology [57], with greater cognitive deficits in severe, long-lasting disorders such as schizophrenia and bipolar disorder [5, 58]. Though in adults, this relationship appears to transcend boundaries of individual psychiatric diagnoses [59], importantly suggesting that cognitive deficits are not confined to specific disorders but rather reflect a broader transdiagnostic phenomenon. This transdiagnostic view is endorsed by child and adolescent clinicians engaging with adolescent patients [60], which points to it being particularly useful for the heterogeneous group of PTB adolescents. The comparable reductions at the beginning and end of adolescence, with a more level cognitive playing field during the height of neural rewiring in mid-adolescence, suggest that cognitive abilities are significantly reduced at two time points with major life changes (i.e., start of adolescence and reaching legal adult age). Importantly, increasing differences from age 15 to 19 may be suggestive of persisting differences going into adulthood [30, 39, 41].

The reviewed literature also shows that PTB adolescents have significant deficits in gross motor skills, particularly balance and coordination [30]. The observed balance deficits continue into adolescence, still being present at age 14 [30]. Cognitive and academic development can be further compromised by gross motor deficits, which have been shown to exacerbate existing cognitive difficulties and hinder academic achievement[61–64]. In addition, motor difficulties have been linked to long-term effects on psychosocial adjustment, emotional well-being, and reduced social engagement [65].

Fine motor impairments are similarly pronounced in PTBs, with adolescents showing lower performance on tasks such as the RCFT [44]. As fine motor skills are intrinsically linked to cognitive performance [65, 66], deficits in fine motor skills can also negatively impact cognitive functioning. Findings of fine motor skills predicting higher academic achievement in subjects like English and science corroborate this link to cognition and these skills relevance [53].

One possible explanation for reduced motor skills and cognitive vulnerabilities in PTB populations is atypical cerebellar development. Xu et al. [67] found greater structural covariance between the cerebellum and frontal and parietal lobes in preterm infants compared to full-term controls, suggesting altered structural connectivity in motor- and cognition-relevant circuits. Complementing this, Wu et al. [68] reported smaller cerebellar hemispheres— particularly in the superior left hemisphere—and enlarged anterior vermis and posteroinferior lobes in preterm infants at term-equivalent age. These structural differences likely stem from disrupted proliferation and differentiation of cerebellar neurons, which typically occurs during the last trimester of pregnancy and is interrupted by preterm birth [69].

Large-scale cohort studies [38], however, suggest that not all PTB populations experience motor deficits. This variability in motor outcomes among PTB individuals may stem from the heterogeneous nature of PTB populations, their individually experienced birth insults and negative factors that studies often do not differentiate between. For more mechanistic insights on the degree of motor difficulties and contributing factors such as gestational age, birth complications, and variations in postnatal care, we recommend to consider these aspects in future studies [70].

While frequent early visual alterations manifesting in cerebral visual impairment (CVI) are known in preterm populations, the limited evidence on visual abilities reviewed herein might have its reasons in excluding publications with participants suffering from CVI. However, the identified studies, analysing PTB adolescents without CVI, demonstrate existing visual deficits that persist beyond childhood [46]. These more subtle deficits in visual perception, visuo-motor control, and visuo-spatial planning do not only remain but become more pronounced later on during adolescence in PTB individuals compared to their FT peers [36, 71]. These findings underline that functional visual abilities can be impacted by PTB, even in the absence of more severe CVI. Generally speaking, accurate visual functioning is assigned importance, since its absence may have an enduring impact on academic performance, daily life, and overall mental health [72, 73]. The fact that visual difficulties may be influenced by atypical neurodevelopment or neurosensory integration [74, 75] links the reviewed domains and underlines the complexity of challenges after PTB.

The present review highlights consistently reduced performance across motor, cognitive, and visual abilities in a considerable number of adolescents after PTB. Given these abilities’ profound role in daily functioning, it is essential to assess these domains comprehensively when considering the long-term health outcomes of PTB individuals. For instance, adolescents’ psychopathology may not be adequately predicted by cognitive outcomes alone. Adding visual assessments, which are often considered secondary to cognitive and psychiatric outcomes [14], may improve prediction further. Including aspects of motor skills, which contribute to cognitive function and daily functioning, may also enhance predictive accuracy— an approach supported by the primacy of vision in neural processing and developmental organization [76]. For instance, removing visual information in healthy sighted individuals temporarily leads to misperceptions such as hallucinations [15]. A more comprehensive approach may also consider the inclusion of genetic risk scores for developing a mental health disorder later in life. These scores are posited by some neurobiological models of psychosis that highlight early prenatal vulnerabilities in the disorder’s development [37, 77]. Hence, assessing all aforementioned factors appears crucial for understanding the full spectrum of challenges faced by PTB individuals many years after neonatal care typically ends. If not addressed, these challenges might result in worse mental health outcomes.

The findings of this review have important implications for clinical care of PTB adolescents. Clinicians should be aware of the interplay between motor, visual, and cognitive abilities and their impact on mental health [78–80]. Systematic screening for visual deficits, even in the absence of CVI, is essential in PTB individuals with social, perceptual, or cognitive difficulties, as vision and motor skills play a key role in development [75, 81].

Given the elevated risk of psychiatric disorders in PTB populations [5, 8, 9, 82], our results suggest that routine mental health screening during early and later adolescence would be beneficial. Many psychiatric conditions emerge or show early signs during this period [5, 7], underlining the need for ongoing developmental monitoring [83].

The investigation of different functional domains presents both challenges and opportunities for future research efforts. Given the elevated risk of PTB individuals to suffer from CVI, scarce data concerning their long-term visual abilities needs expansion. A particular focus could be placed on the triangulation between visual and cognitive aberrancies and their contribution to the increased risk of developing mental ill-health [5, 22]. Longitudinal studies combining cognitive, motor, visual, and neuroimaging data would be invaluable for understanding the long-term trajectories and their mechanisms in adults born PTB [53]. Longitudinal investigations of visuo-motor skills could use eye-tracking technology to offer valuable insights into the cognitive and somatic mechanisms underlying developmental differences [84].

Similarly, investigations of the predictive power of fine and visual-motor coordination as well as genetic and environmental factors as add-ons to current early detection approaches of psychiatric disorders are important future avenues [79, 80]. Their inclusion may lead to better mental health outcomes for PTB populations. Genetics has already revealed a link between altered intrauterine gene expression under stress as a vulnerability for developing severe mental health disorders such as psychosis later in life [37, 77], but research on this link in PTB individuals remains scarce.

This systematic review may be impacted by certain limitations. Firstly, although there is a possibility of selection bias due to the restriction to cohorts born after 1 January 1980, this focus ensures the review does not miss crucial information from longitudinal PTB cohorts who have benefitted from improved neonatal care. Thereby, it remains highly relevant to contemporary clinical and developmental contexts and strengthens the applicability of the conclusions to current PTB populations. Secondly, while heterogeneity in assessment methods and diagnostic criteria across the included studies complicates direct comparisons and more extensive meta-analyses, the review’s systematic methodology, incorporating a small meta-analysis of studies with good quality, enabled the most meaningful integration and synthesis of findings. Thirdly, our meta-analyses of IQ scores show one study reporting two substantially larger effect sizes of reduced IQ scores. This could have biased the outcome of this meta-analysis. However, an exaggeration of the observed IQ reduction is unlikely given the complementary analysis of mean IQ differences, where this study does not stand out.

In conclusion, this systematic review’s search approach focused on multiple abilities and their interplay with mental health to gain a comprehensive understanding of the long-term effects after PTB. We find evidence in the literature for the complex and enduring negative consequences of PTB birth circumstances on cognitive and motor development that can persist into adolescence and even adulthood. These difficulties include substantial reductions in IQ score across studies and adolescence, executive functions, and motor coordination skills, particularly of balance. The identified challenges are linked to broader difficulties in academic performance and mental health. Importantly, PTB adolescents are more predisposed to the development of subsequent psychiatric disorders than their FT-peers, most notably in the domain of ADHD. Gender differences for the prevalence of several disorders are noteworthy. Together, the relevance of mental ill-health in PTB adolescents suggests the implementation of early screening efforts. However, we identified a substantial gap of research examining long-term visual abilities in PTB adolescents. Our findings call for more targeted research on visual abilities and the use of the reviewed domains in tailored early detection approaches to address the specific, and often complex needs of individuals born preterm. Ultimately, this can lay the foundation for improving their quality of life.

## Declarations

### Author contributions statement

**Conceptualisation:** F.S., J.O., W.G., S.B., and L.F.

**Data curation:** F.S., H.S., J.W., N.D., L.H., and L.F.

**Formal analysis:** F.S. and L.F.

**Funding acquisition:** C.A., S.B., and L.F.

**Investigation:** F.S. and L.F.

**Methodology:** F.S., K.R., and L.F.

**Project administration:** K.R., S.B., and L.F.

**Resources:** C.A., S.B., and L.F.

**Software:** F.S. and L.F.

**Supervision:** K.R. and L.F.

**Validation:** F.S.

**Visualisation:** F.S. and L.F.

**Writing - original draft:** F.S. and L.F.

**Writing - review & editing:** F.S., E.G., S.E., C.A., K.R., J.O., W.G., S.B., and L.F.

## Acknowledgements

This work was supported by the University of Lübeck’s Faculty of Medicine’s grant for junior scientists (J15-2024 to L.F.).

## Data availability statement

All data generated or analysed during this study are included in this published article. Additional materials related to the meta-analysis (e.g., extracted data tables, risk of bias assessments, and supplementary analyses) are available via the Open Science Framework repository: https://osf.io/me2d6/.

